# Interventions to reduce peripheral blood culture contamination in acute care settings: A systematic review and meta-analysis

**DOI:** 10.1101/2023.07.26.23293230

**Authors:** James A. Hughes, C.J. Cabilan, Julian Williams, Mercedes Ray, Fiona Coyer

## Abstract

**Background:** Blood culture contamination is a significant problem in acute care settings. Contamination of a blood sample with pathogens not present in the patient’s blood leads to increases in length of stay, overuse of antimicrobials, and increases in healthcare cost. Several interventions have been reported in different settings within the literature to decrease the contamination. However, their overall effectiveness is currently unknown.

**Objective:** This systematic review aimed to identify interventions to reduce contamination from peripherally collected blood cultures and to evaluate the effectiveness of these interventions.

**Design:** Systematic review and meta-analysis

**Methods:** In March 2019 we performed a systematic search of English language literature from academic databases, registers of clinical trials and grey literature for interventions aimed at reducing blood culture contamination in adult acute care settings. Studies meeting inclusion criteria were reviewed and data were extracted by two independent reviewers.

**Results:** A total of 6,302 articles were retrieved from searches. After removal of duplicates and screening against inclusion criteria 57 studies were included. The majority of the 57 studies had a medium to high risk of bias. These studies identified eight specific interventions (collection packs, dedicated collection teams, education, staff feedback, intervention bundle, sterile procedure, Initial Specimen Diversion Devices, or change of asepsis solution) used in acute care. Thirty-four studies were included in the meta-analysis. There was a wide variation in the definition of contamination which precluded many studies from being included in the meta-analysis. Dedicated collection teams (RR 0.40, 95%CI 0.21 – 0.76, I^2^ 87%, p<0.001) and initial specimen diversion devices (RR 0.43, 95%CI0.31 – 0.58, I^2^ 84%, p<0.001) were the most successful at reducing blood culture contamination. Heterogeneity was high across all studies and interventions.

**Conclusions:** The use of dedicated collection teams or initial specimen diversion devices showed the most significant reduction in blood culture contamination; however, other interventions such as intervention bundles, education or feedback, may have benefits in terms of ease of implementation, and have still been shown to lower blood culture contamination.

## Introduction / Background

The contamination of blood culture samples with organisms not present in the blood of patients is a significant clinical issue in acute care settings (1). Blood culture contamination is defined by the American Clinical and Laboratory Standards Institute as *“microorganism isolated from a blood culture during specimen collection or processing [and was] not pathogenic for the patient from whom the blood was collected*” ((2)p5). Blood culture contamination leads to unnecessary treatment, extends hospital length of stay and contributes to antimicrobial resistance. Current recommendations support an upper limit of 3% for potential contamination of all blood cultures collected in acute care settings (1–4). However, in practice, figures below 3% are rarely reported, and contamination rates up to 12% are prevalent (1, 5–10).

In North American studies, blood culture contamination has been identified as contributing up to an additional 4.5 days to the average patient length of hospital stay (9, 11). In the United Kingdom, this length of stay is even further extended to 5.4 days (7). Blood culture contamination has been associated with increased use of antimicrobials, particularly vancomycin, which in turn is associated with increased costs of pharmacokinetic monitoring (12). Blood culture contamination also creates a financial burden on health care systems contributing up to $7,500 per patient compared to patients with true negative cultures (1, 7, 9–11).

Blood cultures can be collected through either peripheral venepuncture or central venous access device. Most commonly, blood cultures are collected through peripheral venipuncture or a newly established peripheral intravenous catheter (PIVC). Blood cultures can also be collected through venous access devices such as a Port-a-cath® (Smith’s Medical), Peripherally Inserted Central Catheter (PICC), and Hickmann® (Bard Access Systems) lines.

Several interventions have been reported targeting reductions in blood culture contamination; these have included educational interventions (6, 13), feedback (9), bundled interventions (13), changes to skin asepsis, and specimen diversion devices (5). However, it is not currently known what the grouped effectiveness of these interventions or their effectiveness in different acute care settings such as the emergency department or ward environment. This systematic review aimed to identify interventions to reduce contamination from peripherally collected blood cultures and to evaluate the effectiveness of these interventions.

## Methods

A detailed description of the methods used in this systematic review and meta-analysis have been reported in a previously published protocol (14). The results of the systematic review are reported here according to the PRISMA guidelines (15).

### Data Sources and Search Strategy

A comprehensive search was conducted from inception to March 2019 in: Cumulative Index Of Nursing And Allied Health Literature (CINAHL via Ebsco), Pubmed, Excerpta Medica database (EMBASE), and Proquest Dissertation and Thesis. Controlled trials were searched for via the Cochrane Central Register of Controlled Trials (CENTRAL) from 1996 to March 2019. Search strategy and results for each database are detailed in Table 1.

**Table 1.**
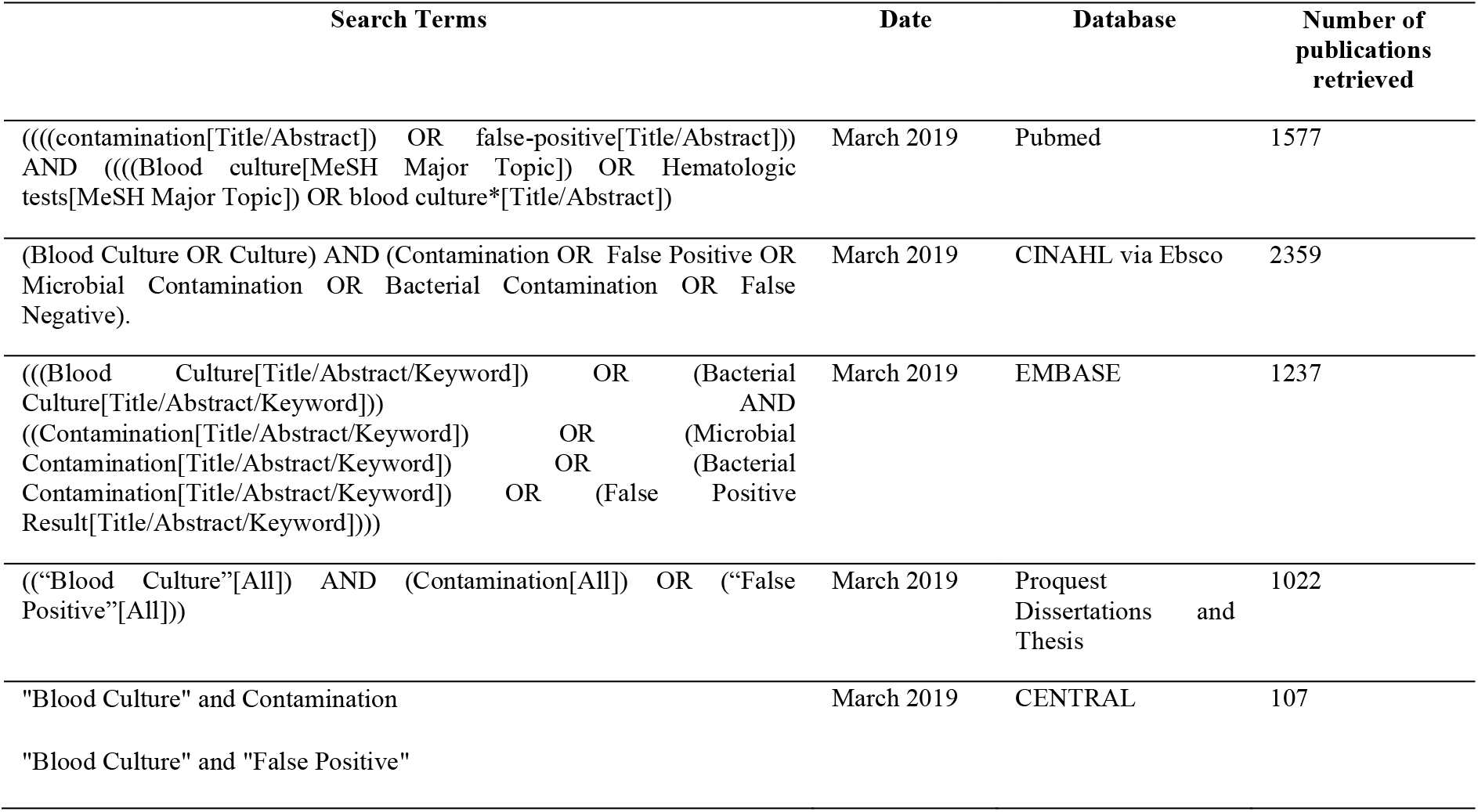
Search strategies for each database searched. CINAHL = Culmulative Index of Nursing and Allied Health Literature, EMBASE = Excerpta Medica database, CENTRAL = Cochrane Central Register of Controlled Trials.

### Inclusion Criteria

Experimental studies (randomised control trials or quasi-experimental studies) that assessed the effectiveness of an intervention or interventions aimed at reduction of blood culture contamination were included. Interventions targeted at adult patients (greater than 16 years old) and cultures collected in acute healthcare (hospital) settings were eligible for inclusion. Secondary outcomes such as multiple sets, volume, the proportion of positive samples and costs were summarised in the results. Due to the lack of funding for translation services, only studies reported in English or with a previous translation available were included in this review.

### Study selection and data extraction

Selection of studies was conducted independently by two reviewers (JAH and CJC). Discordant assessments were discussed between the two reviewers without the need for review by a third reviewer (FC). All citations from initial searches were imported to a reference library and screened for relevance using titles and abstracts. Full texts of all relevant citations were accessed and subsequently screened against the inclusion criteria. The methodological quality of each selected study was appraised using the Effective Practice and Organisation of Care group risk of bias criteria (16).

### Meta-analysis

Data extraction from selected studies was conducted and cross-checked independently by two reviewers (JAH and CJC) for accuracy. Data were entered into Review Manager (RevMan, version 5.4) software for meta-analysis (17). The primary outcome (blood culture contamination rate) was analysed using the Mantel-Haenszel method within a random-effects model. Risk ratios (presented with 95% confidence intervals) of less than one represented a reduced contamination rate for the intervention. Heterogeneity was assessed using the I^2^ statistic and considered significant at the p=0.1 level (18).

Consensus was reached to group interventions as: dedicated blood culture collection teams, intervention bundle including collection kits, education, initial specimen diversion device, staff feedback, sterile procedure. Studies were only included in the meta-analysis if they had a clear definition of contamination that included at least three of the seven most commonly reported contaminants (19):

- *Coagulase-negative staphylococci*
- *Viridans streptococci*
- *Propionibacterium spp*.
- *Micrococcus spp*.
- *Corynebacterium spp*.
- *Diphtheroids*
- *Bacillus spp*.

## Results

A total of 6302 articles were identified from the searches. After removal of duplicates, 5065 articles were screened for relevance, which subsequently reduced the number of articles to 175 for screening against the inclusion criteria (Figure 1), of these 57 were included in the systematic review. Characteristics of included studies can be found in Table 2. The majority of studies (n=37, 65%) were quasi-experimental, either pre and post-test design or a time series (6, 8, 9, 13, 20–52). Sixteen studies (28%) used an experimental design (5, 53–67), and four (7%) used other studies designs (1, 68–70). Thirty-four studies included sufficient data for quantitative synthesis in the meta-analysis (5, 6, 9, 20, 22–25, 28–30, 33, 34, 36–38, 44, 45, 47–50, 52, 55–59, 61, 62, 65–67, 70).

**Figure 1:**
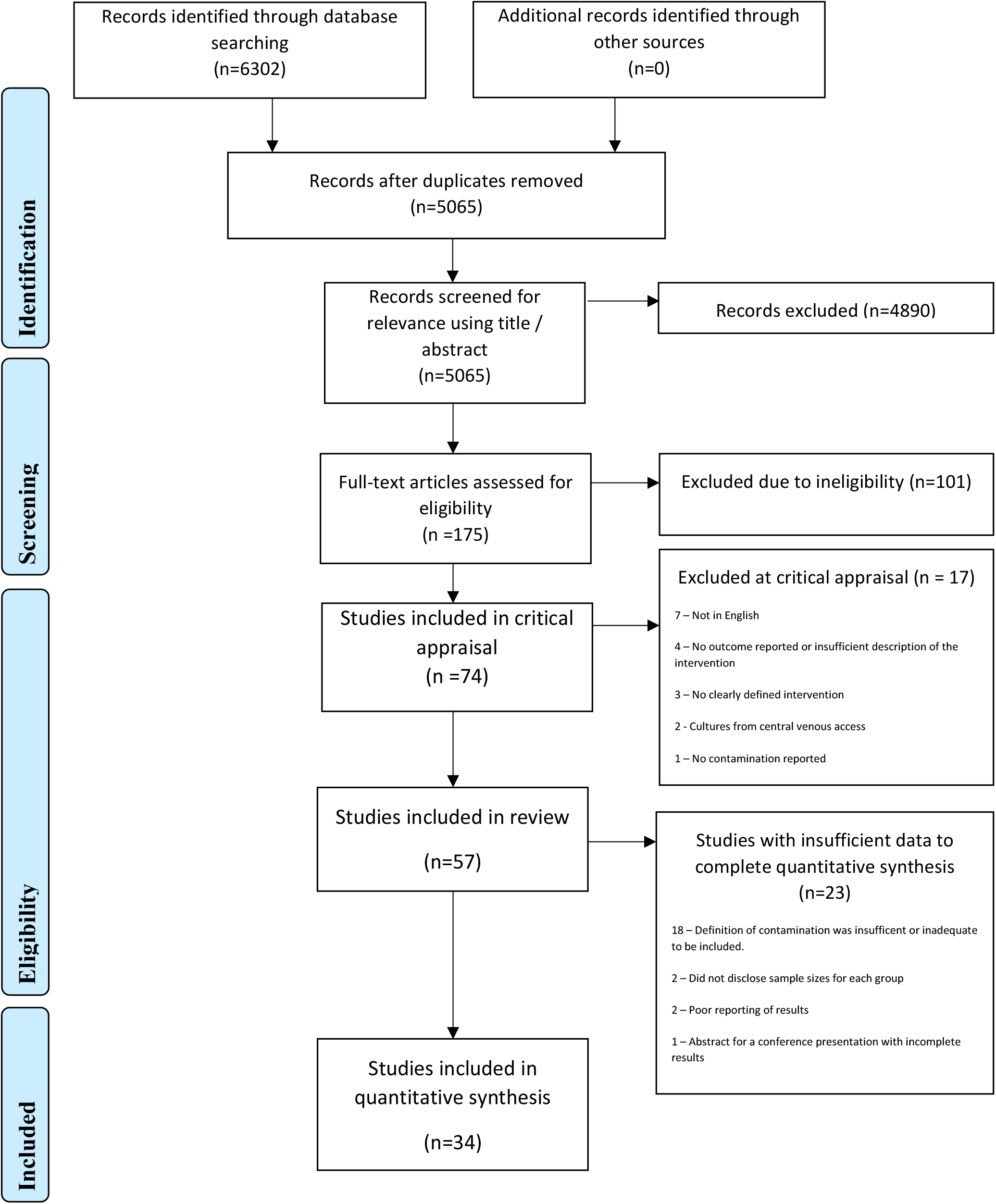
Systematic review study (PRISMA) flow diagram (15)

**Table 2:**
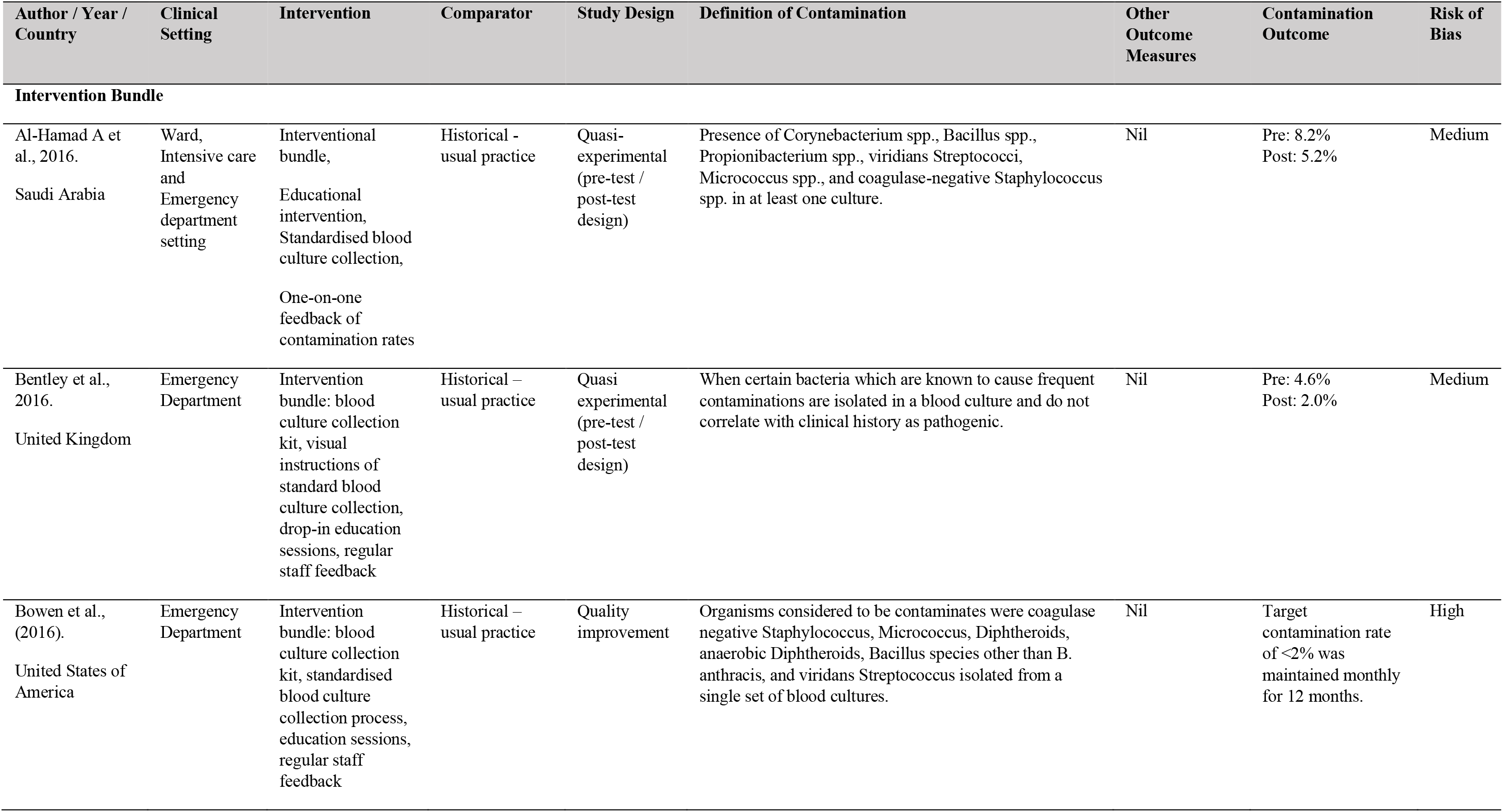
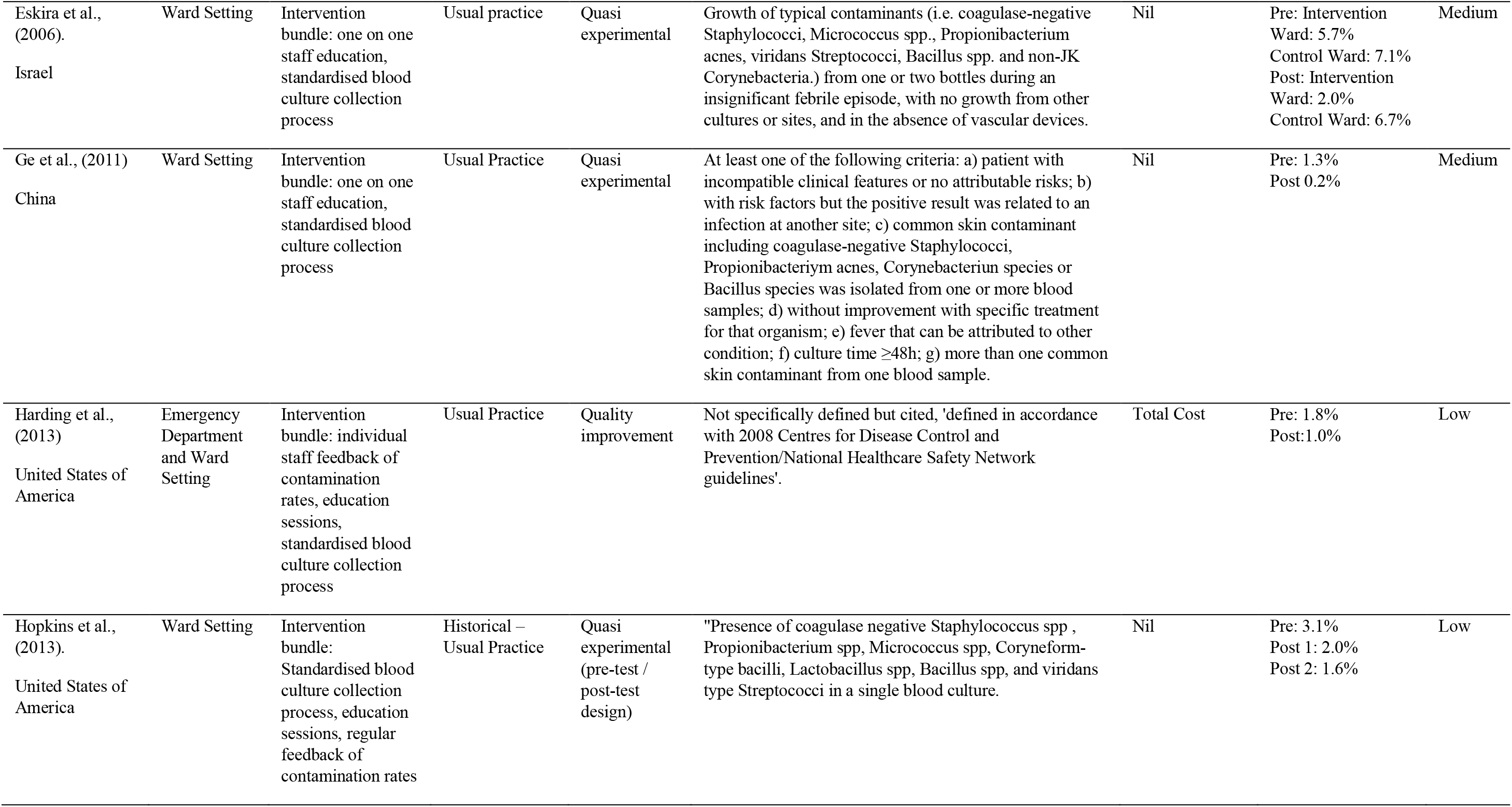
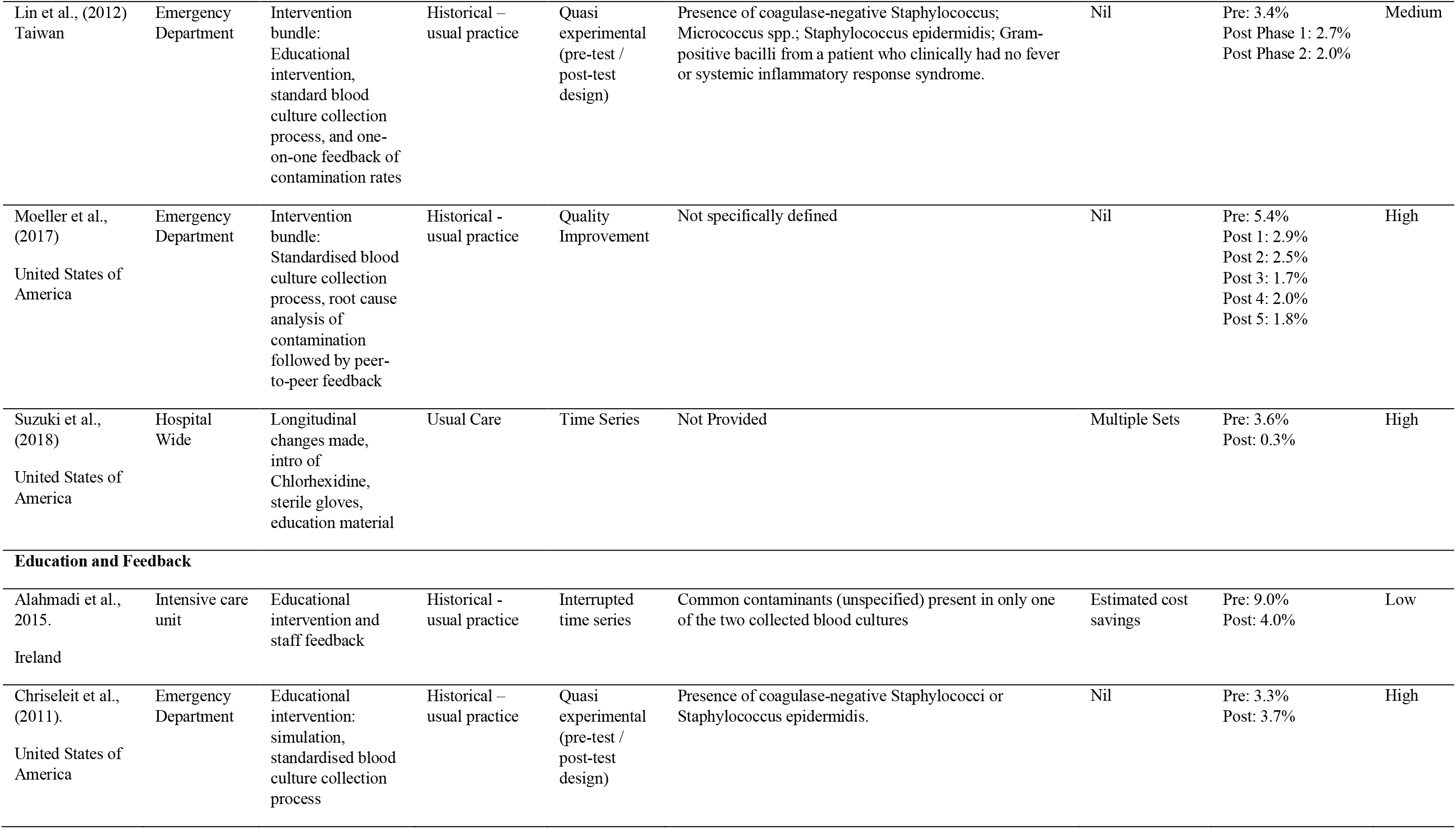
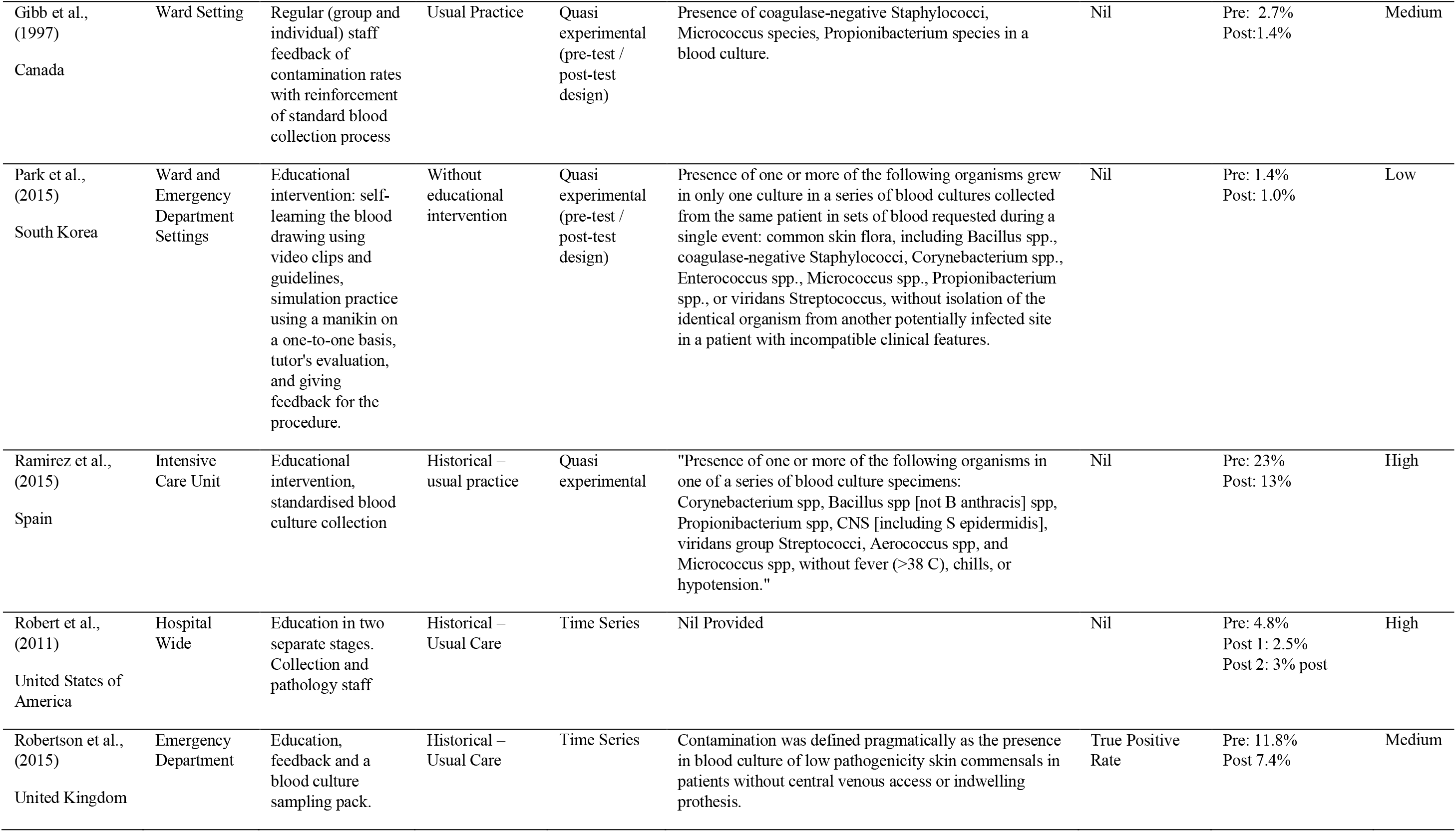
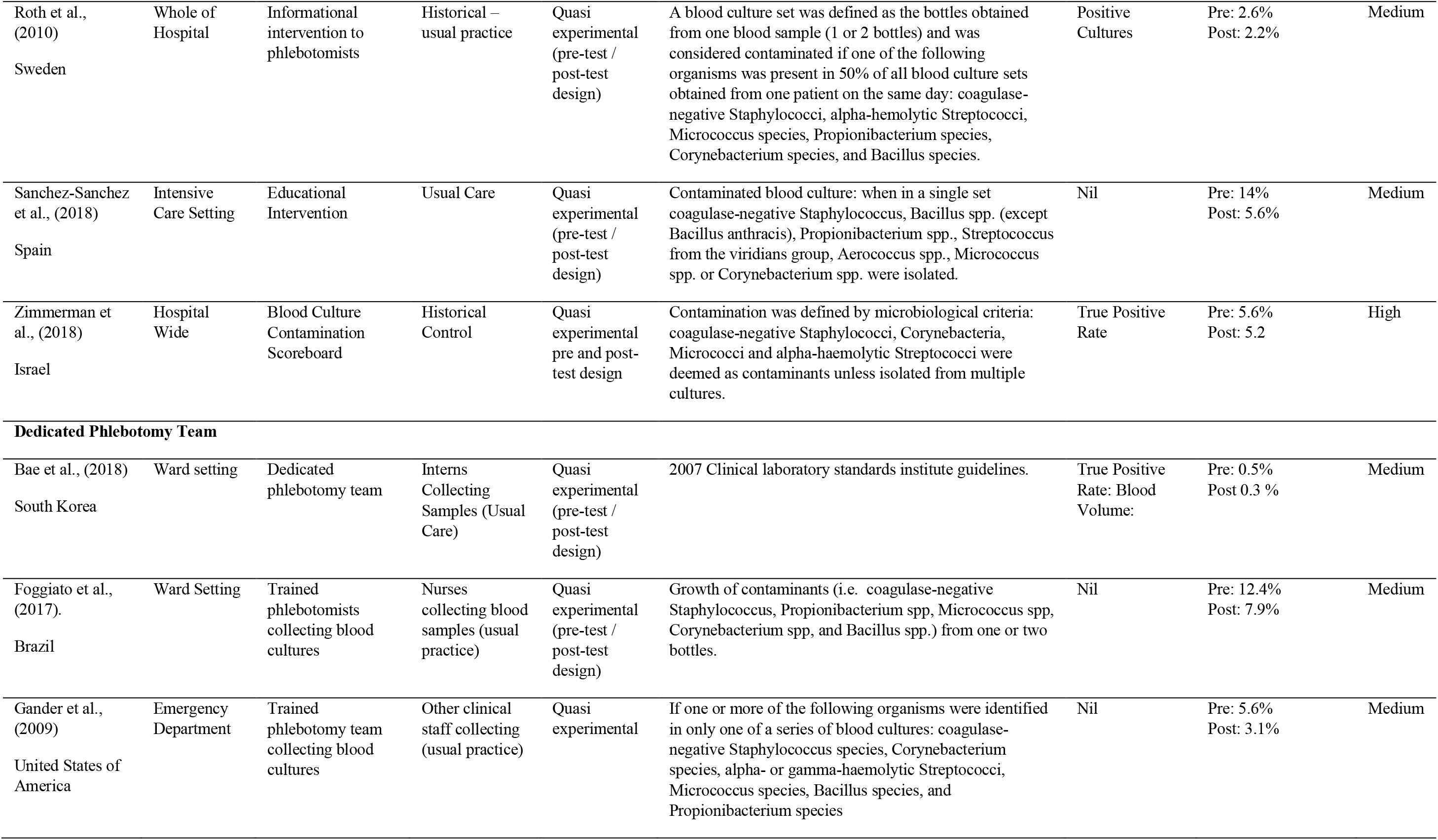
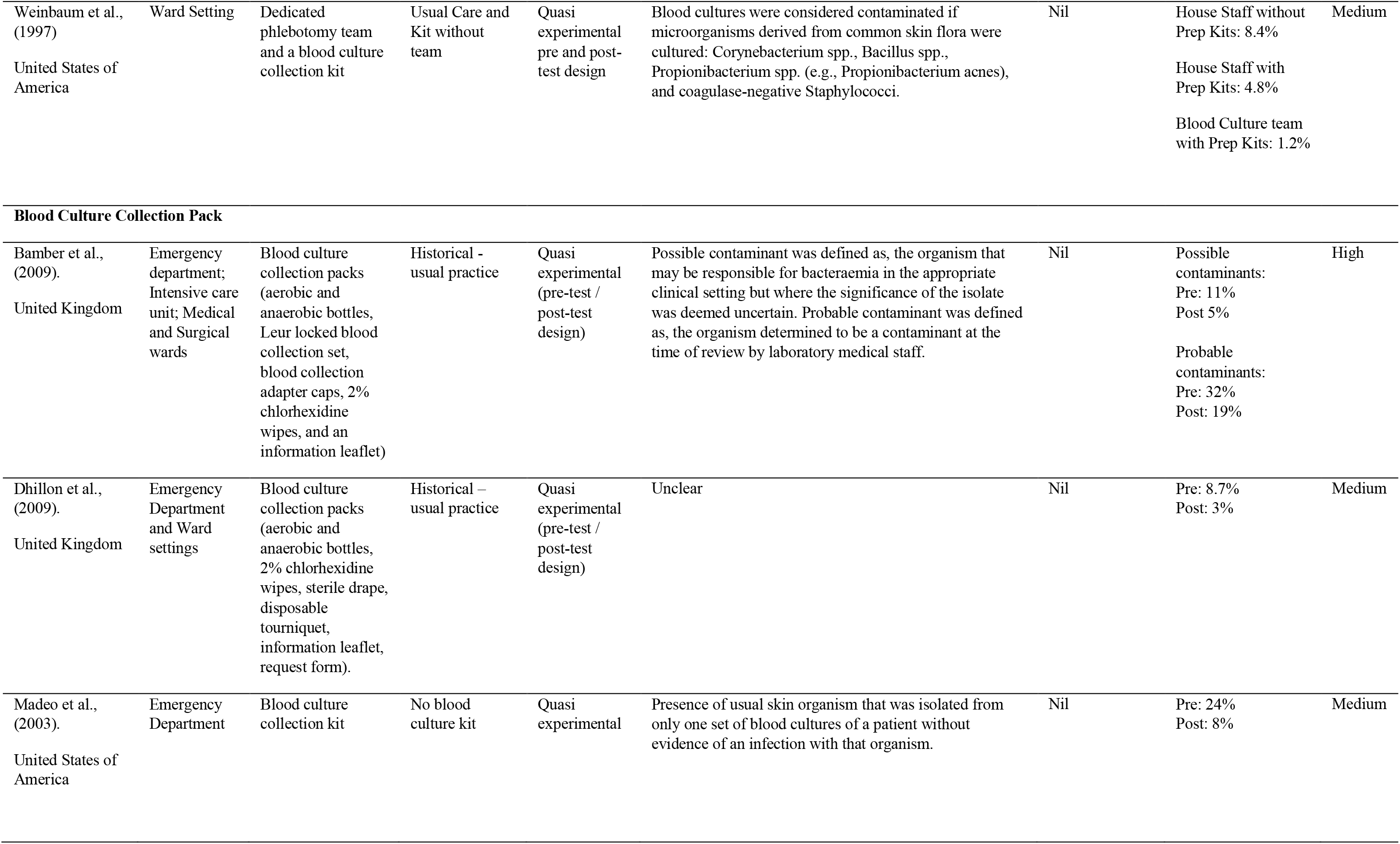
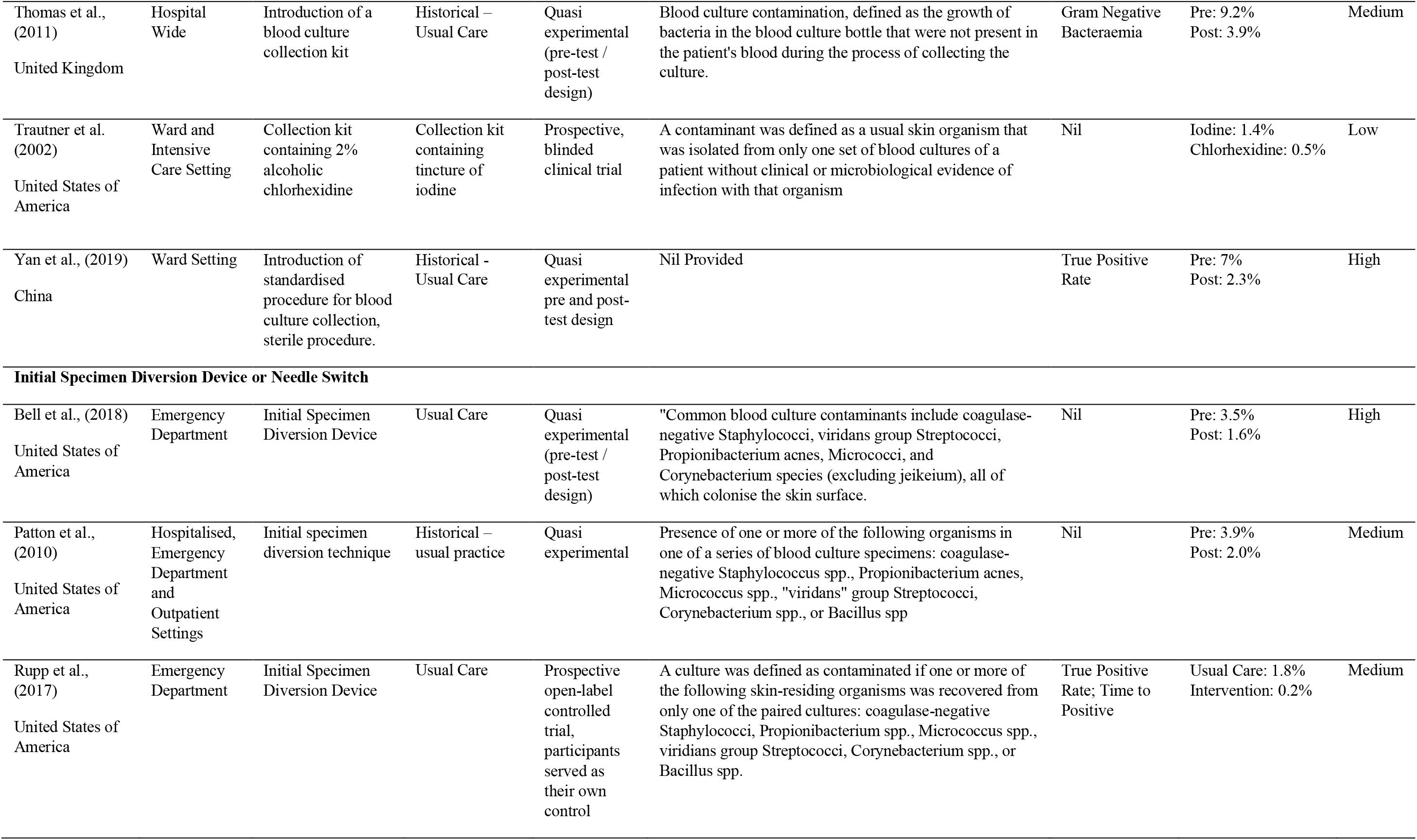
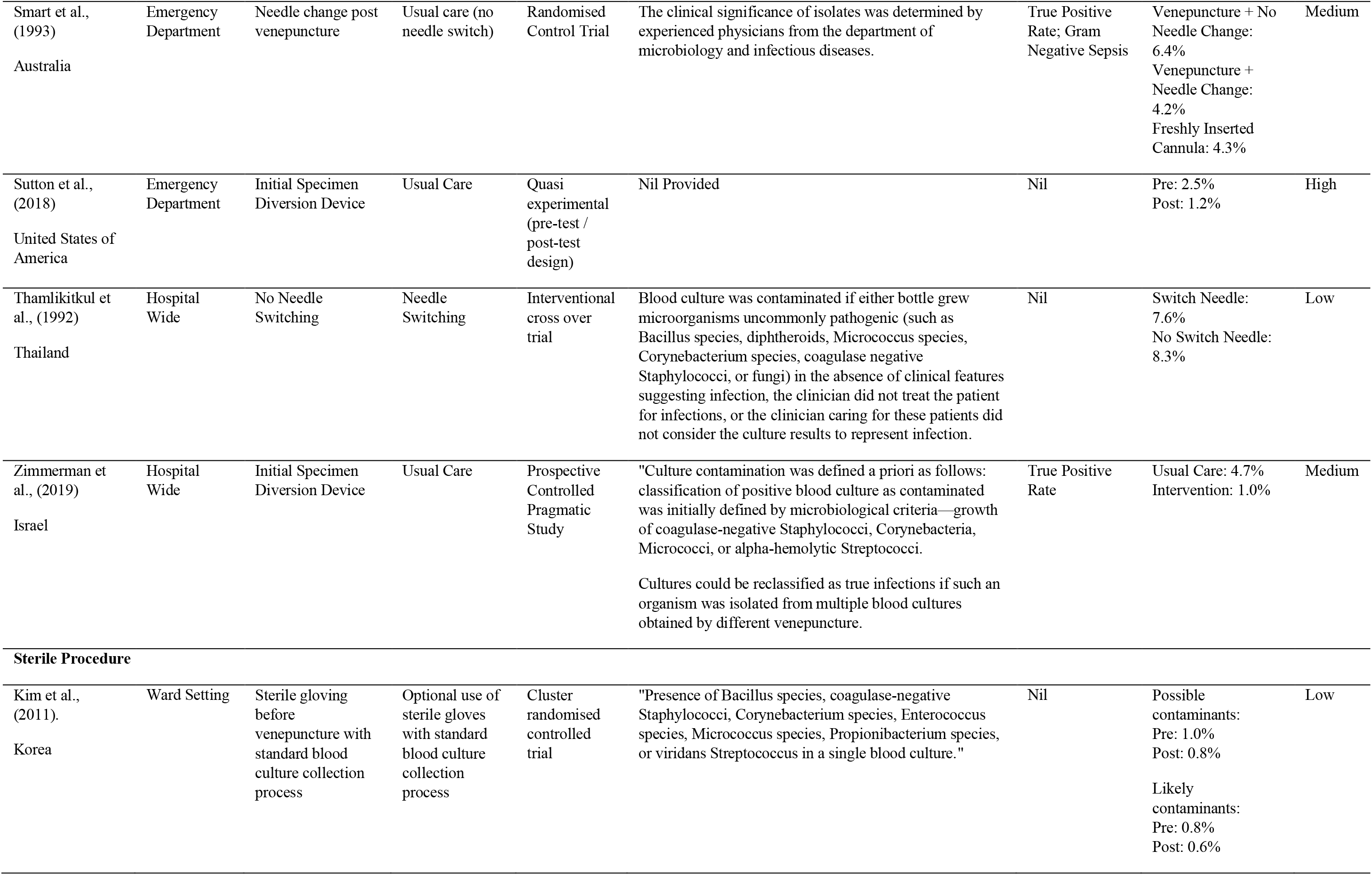
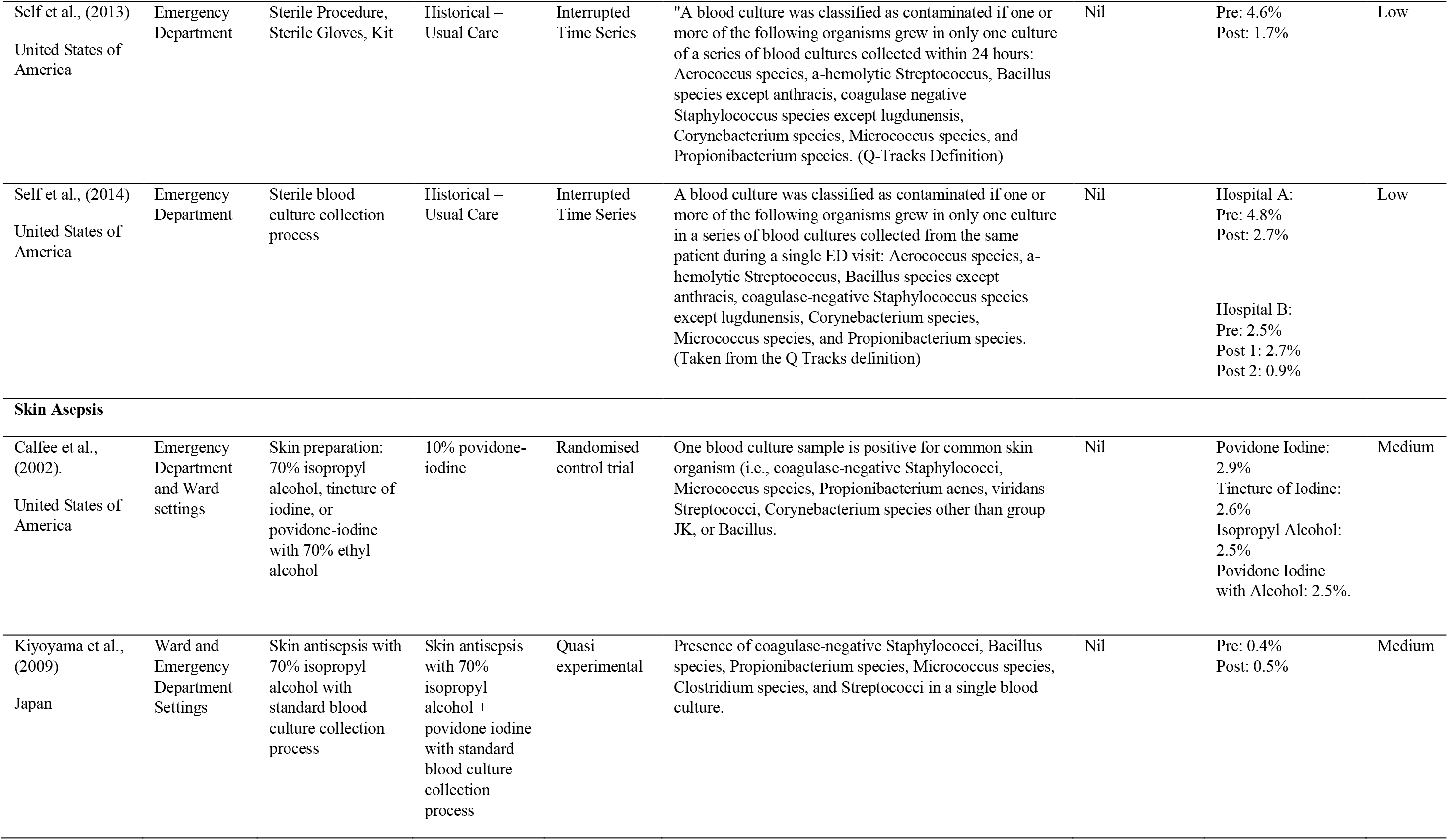
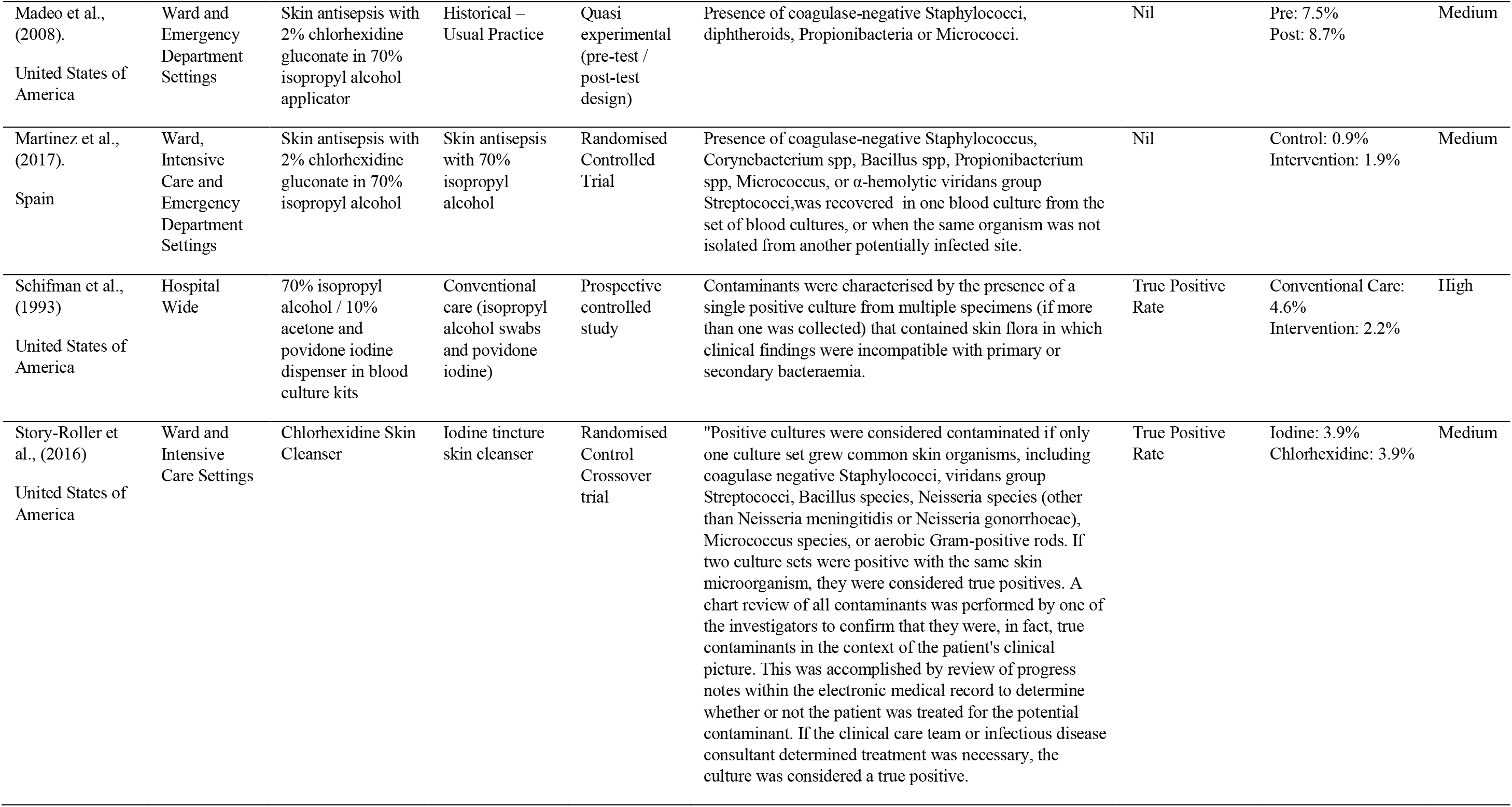
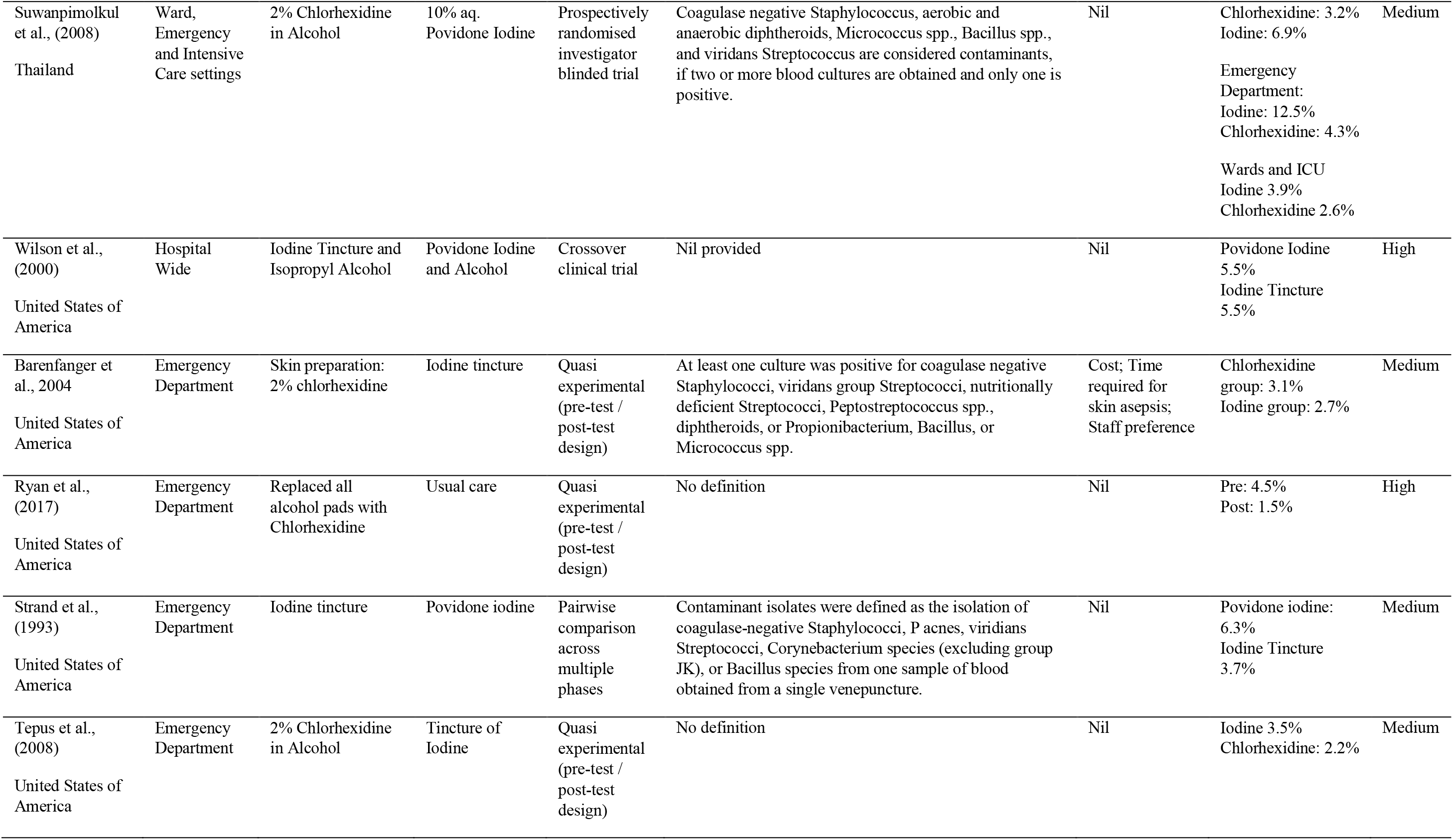
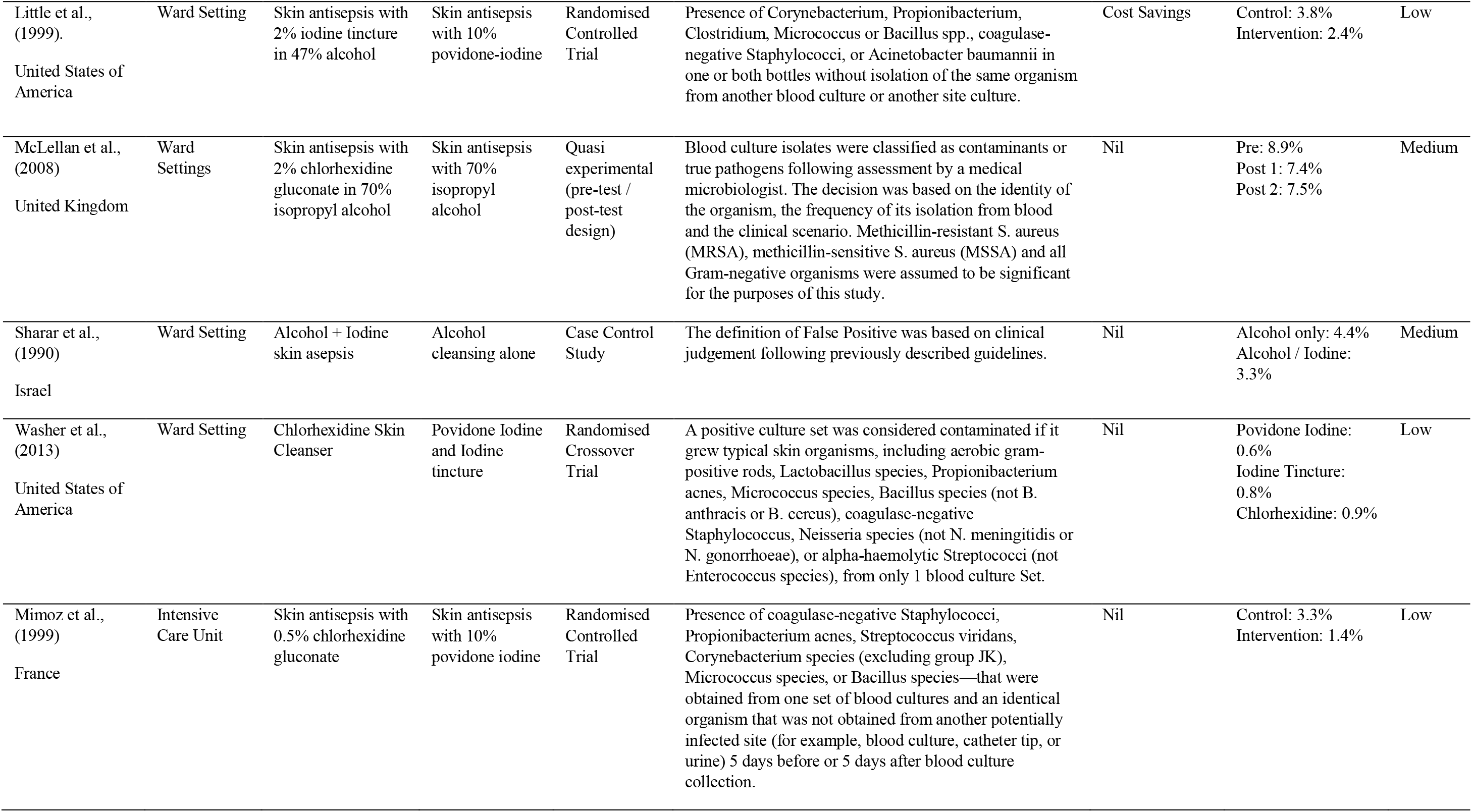
Summary of included studies.

### Settings

From the studies included, 18 (31.6%) were conducted in the emergency department (1, 5, 8, 9, 13, 21, 25, 32, 37, 41, 44, 46, 47, 50, 51, 70–72), 13 (22.8%) were conducted in ward environments (22–24, 28, 35, 36, 38, 43, 57, 64, 65, 67, 73) and four (7.0%) were conducted in the intensive care unit setting (27, 30, 33, 66). Combinations of these areas accounted for 13 (22.8%) of studies (20, 29, 39, 40, 45, 48, 53, 58, 59, 61, 68, 74, 75), and nine (15.8%) studies were described as hospital-wide (6, 26, 31, 34, 42, 55, 56, 60, 63).

### Quality and Bias

The risk of bias was overall was assessed as high in 13 (22.8%%), and medium in 32 (56.1%) of the studies included in this review. The high level of bias is reflective of the quasi-experimental methodology of the majority of the studies with a lack of randomisation procedures, or poorly reported randomisation. Differences in baseline characteristics were either not reported or not taken into consideration in a number of the included studies. A total of 12 studies (21.1%) were assessed as low risk of bias. The risk of bias for each included study is summarised in Supplementary Tables 1 and 2.

### Interventions

Interventions to decrease the rate of blood culture contamination varied widely throughout the included studies. Most studies used a combination of initiatives aimed at intervening at different points in the collection process where contamination can occur. Eight distinct types of interventions were identified: education delivered to clinicians who collect blood cultures (6, 13, 20–27, 29–33, 68, 76), staff feedback (20, 21, 24, 25, 27–29, 32, 34, 68, 69, 76), dedicated phlebotomy team to collect blood cultures (36–38, 77); blood culture collection packs containing all necessary equipment (21, 32, 38–42, 47, 53, 60, 76), intervention bundles with standardised blood culture collection processes (13, 20–25, 28, 30, 40, 43, 48, 57, 68, 69, 76), a change to the devices used such as a needle switch or initial specimen diversion devices (44–46, 55, 56, 78) or the introduction of sterile procedures (26, 47, 57, 79). Many of the studies used a combination of these interventions in an attempt to decrease contamination rates. Full details of each study can be seen in Table 2.

The most common intervention to decrease contamination in blood cultures was skin antisepsis before venepuncture (8, 48, 50, 51, 53, 58–61, 63–67, 70, 73–75). A wide variety of antiseptic solutions were used as either “usual care” or as an intervention. For example, chlorhexidine, iodine and alcohol preparations were used as either “usual care” or as an intervention. Table 3 shows the skin asepsis solutions used as intervention or usual care and the contamination rates found in each study.

**Table 3:** Studies investigating the change of skin asepsis solutions on contamination rates. ED = Emergency Department; W = Ward; ICU = Intensive Care Unit; H = Whole of Hospital; MO = Medical Officer; RN = Registered Nurse; P = Phlebotomist; T = Health Care Technician

| Author / Year | Sample Size | Setting | Clinician | Skin Asepsis (Intervention) | Contamination | Skin Asepsis (Usual Care) | Contamination |
| --- | --- | --- | --- | --- | --- | --- | --- |
| Barenfanger et al., (2004) | 11738 | ED | MO & RN & P | 2% chlorhexidine | 5% | Iodine tincture | 11% |
| Calfee et al., (2002) | 12692 | ED & W | Not Specified | 70% isopropyl alcohol | 2.50% | 10% povidone-iodine | 2.93% |
|  |  |  |  | Tincture of iodine | 2.58% |  |  |
|  |  |  |  | Povidone-iodine with 70% ethyl alcohol | 2.46% |  |  |
| Kiyoyama et al., (2009) | 5653 | ED & W | MO | 70% isopropyl alcohol | 0.42% | 70% isopropyl alcohol + povidone iodine | 0.46% |
| Little et al., (1999) | 3851 | W | P | 2% iodine tincture in 47% alcohol | 2.42% | 10% povidone-iodine | 3.8% |
| Madeo et al., (2008) | 100 | ED & W | MO | 2% chlorhexidine | 2.1% | Not defined | 7.5% |
| Martinez et al., (2017) | 1102 | ED & ICU & W | RN | 2% chlorhexidine | 1.86% | 70% isopropyl alcohol | 0.89% |
| McLellan et al., (2008) | 1725 | W | MO & T | 2% chlorhexidine | 7.43% and 7.54% | 70% isopropyl alcohol | 8.88% |
| Mimoz et al., (1999) | 2041 | ICU | RN | 0.5% chlorhexidine | 1.37% | 10% povidone iodine | 3.33% |
| Ryan et al., (2017) | 378 | ED | MO & RN | 3.15% chlorhexidine | 1.5% | 70% isopropyl alcohol | 4.5% |
| Schifman et al., (1993) | 1546 | H | MO & RN | 70% isopropyl alcohol / 10% acetone / povidone iodine (PREP method) | 2.2% | Isopropyl alcohol and povidone-iodine | 4.6% |
| Shahar et al., (1990) | 362 | W | MO | 70% isopropyl alcohol and providone iodine solution applicator | 3.3% | 70% isopropyl alcohol | 4.4% |
| Story-roller et al., (2016) | 6095 | ICU & W | RN & T | 2% chlorhexidine | 3.88% | 70% isopropyl alcohol and iodine tincture | 3.93% |
| Strand et al., (1993) | 12795 | ED | MO & RN | Iodine tincture | 3.74% | Povidone-iodine | 3.74% |
| Suwanpimolkul et al., (2008) | 2142 | ED & ICU & W | MO & RN | 2% chlorhexidine | 3.2% | Povidone-iodine | 6.25% |
| Tepus et al., (2008) | 14764 | ED | RN & T | 2% chlorhexidine | 2.2% | Iodine tincture | 3.5% |
| Trautner et al., (2002) | 430 | ICU & W | MO & T | 2% chlorhexidine | 0.5% | Iodine tincture | 1.4% |
| Washer et al., (2013) | 8674 | W | P | 2% chlorhexidine | 0.93% | Povidone iodine | 0.58% |
|  |  |  |  |  |  | Iodine tincture | 0.76% |
| Wilson et al., (2000) | 12367 | H | MO | Iodine tincture | 5.5% | Povidone iodine | 5.5% |

### Primary outcome

#### Definition of contamination

The definition of contamination varied widely amongst the studies included in this review. Nine (15.8%) (26, 31, 40, 43, 46, 51, 63, 69, 80) studies did not define contamination at all. The majority (34, 59.6%) of studies used species-based criteria for the definition of contamination with provisions that a contaminant could be considered a true pathogen if it was found in multiple cultures (6, 13, 20, 22, 24, 25, 28–30, 33, 34, 36–38, 44, 45, 47, 48, 50, 55–59, 61, 65–67, 70, 74–76, 78, 79). The remaining 14 (24.6%) studies defined a contaminant without stating a specific species, but used expert opinion or referred to previously published definitions of contamination (21, 23, 27, 32, 35, 39, 41, 42, 53, 54, 60, 64, 68, 73).

#### Meta-analysis

Thirty-four studies were included in the meta-analysis. The remaining 23 studies did not satisfy the review’s definition of contamination, or included insufficient detail in the reporting of their results. Figure 2 displays the results of the meta-analysis by intervention type.

**Figure 2:**
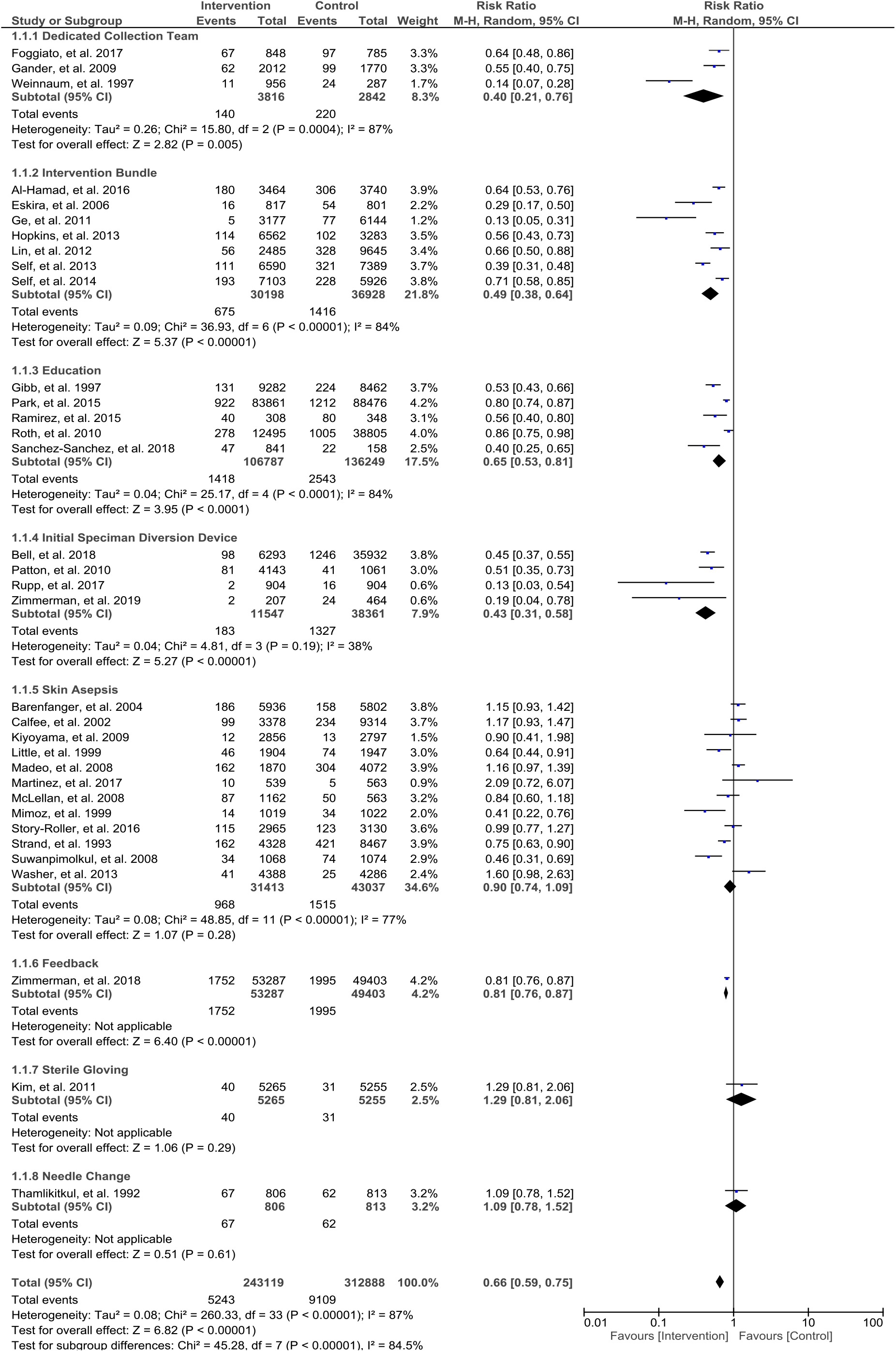
Forest plot of the studies included in the meta-analysis based on intervention. M-H = Mantel-Haenszel, CI = Confidence Interval.

In the 34 studies (N=556,007) included in the meta-analysis, there were eight distinct interventions reported (dedicated collection team; intervention bundle including collection kit, education, initial specimen diversion device, skin asepsis; feedback, sterile gloving, or needle change). Overall, interventions to reduce blood culture contamination significantly decrease the risk of blood culture contamination by 34% (RR 0.66, 95% CI 0.59-0.75; I^2^ 87%) (Figure 2). Heterogeneity was significantly high for all interventions, which could be attributed to the differences in interventions and variation in settings and patient acuity.

Skin antisepsis was the most commonly investigated intervention, but its impact on blood culture contamination is inconclusive. The most impactful intervention appeared to be having a dedicated phlebotomy team for blood culture collection, which resulted to a reduction of blood culture contamination by 60% (RR 0.40, 95% CI 0.21-0.76; I^2^ 87%). This was followed by diversion devices that led to a 57% reduction of contamination (RR 0.43, 95% CI 0.31-0.58; I^2^ 38%). Intervention bundles, usually a combination of education, feedback, collection packs and change of cleaning solution, were associated with a 51% reduction of contamination (RR 0.49, 95% CI 0.38-0.64; I^2^ 84%). Staff education and training led to a 36% reduction of contamination (RR 0.64, 95% CI 0.53-0.81; I^2^ 84%) (see Figure 2).

### Other outcomes

There were eight other outcome measures reported in 57 studies: true positive rate (n=10, 17.5%) (6, 32, 34, 35, 43, 54, 56, 60, 61, 78); cost savings (n=4, 7%) (27, 50, 67, 68), gram-negative isolates (n=2, 3.5%) (42, 54), blood volume (35), staff preference (50), time required for skin asepsis (50), time to positive (78) and multiple sets (26).

Seven of the studies that reported on true positive rates did not detect a change between control groups and the intervention group (6, 32, 43, 56, 60, 61, 78), however these interventions were varied. There were three studies that showed improvements in true positive rates. Bae et al. (2018) showed that the introduction of a dedicated phlebotomy team increased the true positive rate (5.87% vs 5.01%) (77). Smart et al. (1993) showed that the true positive rate increased when cultures were collected from a pre-existing IVC (54). Zimmerman et al. (2018) showed a decrease in the true positive rate of blood cultures during their information/feedback intervention from 5.6% to 5.2% (34). Due to the large sample size of this study, this decrease was significant. However, the clinical significance of a 0.4% decrease in true positive rates is unknown (34). Gram negative isolates fell by 36% post the introduction of a blood culture collection kit, however this did not change the probability of a gram-negative isolate (42). In earlier work Gram negative isolates were higher when blood taken through an existing intravenous access device however did not change through the needle switch intervention (54).

Blood volume cultured was measured by Bae et al., (2019). During their intervention, where the collectors were changed from intern medical officers to a dedicated phlembotomy team, the volume of blood cultured increased from 2.1mls to 5.6mls (35). This increase in volume was associated with a signifigant decrease in contamination rates from (0.45% to 0.27%, p<0.001) (35). The number of sets of blood cultures collected were measured by Suzuki et al., (2018) as part of thie educational intervention. An increase from of multiple sets from 51% to 95% was associated with a reduction of the contamination rate from 3.6% to 0.3% (p=0.012) (26).

In the studies that reported costsavings the results varied widely. Alshmadi et al. (2015) and Harding et al. (2013) reported annualised savings credited to blood culture contamination reduction of £250 000 and $614 000 respectively (27, 68). Other cost saving have been attributed to a per patient saving of $4100 (67) and a saving in asepsis solution of 16c per applicator (50).

## Discussion

This systematic review identified 57 studies that aimed to decrease peripheral blood culture contamination in acute care settings. These 57 studies utilised eight specific interventions in four distinct acute care settings. In general, the interventions reported were found to reduce the risk of blood culture contamination. However, dedicated collection teams, initial specimen diversion devices, intervention bundles or staff education and training were the most successful at reducing contamination. The majority of studies included in this review were quasi-experimental, and without randomisation, therefore their risk of bias was medium to high, with only 12 (21.1%) having a low risk of bias.

Dedicated collection teams have demonstrated the most significant success in reducing blood culture contamination. The introduction of a dedicated team to collect blood cultures has a high cost to the organisation, which limits the ability of organisations to implement this intervention. The introduction of an initial specimen diversion device was also very successful at reducing blood culture contamination. This intervention first appeared in the literature in 2010 and consisted of a diversion of a quantity of blood into a collection tube before collecting the culture samples (45). Since then, authors have described a specifically designed collection device that automatically diverts 1.5 – 2 mLs of blood before collection of the culture sample (5, 44, 56). While this intervention was successful, it also requires specific equipment (at a cost) and training for staff which may not be practical in all settings. Other successful interventions such as, intervention bundles, staff education and staff feedback are also successful methods for blood culture contamination reduction and may have a lower cost to the organisation.

The reduction in blood culture contamination seen by Self et al. (47) when introducing a sterile procedure was not seen in the study by Kim et al. (57) however the difference in approach to the sterile procedure may account for the differences seen. Multiple different skin asepsis solutions have been tried in an attempt to reduce contamination (Table 3). The comparators for the introduction of Chlorhexidine were varied in type (iodine based preparations, alcohols, acetone or combination of these substances) and also varied in terms of the application method. All of these solutions can provide skin asepsis; however, their use and effectiveness vary depending on their application. Differences in the outcomes of these studies may be due to the nature of the application of the substance rather than the substance itself. For these reasons, these results must be interpreted with caution.

Clear trends are present in successful interventions. Education and feedback to staff that collect blood culture, whether this is dedicated staff or general clinical staff have a dramatic impact on the contamination rate. A bundled approach that contains correct equipment, accompanying education and feedback are likely to be deliverable in most settings without the need for dedicated teams or the introduction of new equipment such as initial specimen diversion devices. However, there are benefits in the reduction of blood culture contamination that may necessitate the use of dedicated collection teams and initial specimen diversion devices in settings where reduction can not be achieved by education, feedback and bundled interventions alone. There was no support in the studies included in this review for needle swapping or a sterile procedure.

### Limitations

Systematic reviews and their associated meta-analyses are reliant on the quality of the original studies they incorporate. Overall in this review, many of the studies had a high risk of bias. Therefore there is an increased risk that the overall results contain bias. Some studies with large sample and effect sizes may skew the results. The heterogeneity of the definitions of contamination and methodology used to assess contamination will lead to differences in the reporting of results. Many of the studies reported used multiple interventions simultaneously, which makes it difficult to discern the effect of each intervention; however, in a multifactorial problem such as blood culture contamination this may be a positive attribute. Incomplete data was reported in some studies which may introduce bias into the analysis presented.

## Conclusions

Eight groups of interventions were identified in this review aimed at reducing blood culture contamination in acute care. Dedicated phlebotomy teams, diversion devices, intervention bundles, and staff education and training led to significant reductions in blood culture contamination.

## Data Availability

All data produced in the present work are contained in the manuscript

## Acknowledgements

JAH was supported by a capacity-building grant from the Emergency Medicine Foundation (EMCB-402R23-2015) during part of this work.

Following guidelines outlined in the PRISMA-P methodology, this protocol was registered with the International Prospective Register of Systematic Reviews (PROSPERO) on 8th December 2017, and last updated on 29^th^ January 2020 (registration number CRD42017081650).

## Notes

### Competing Interest Statement

The authors have declared no competing interest.

